# Development of a saliva-optimized RT-LAMP assay for SARS-CoV-2

**DOI:** 10.1101/2020.12.26.20248880

**Authors:** Albert Dayuan Yu, Kristina Galatsis, Jian Zheng, Jasmine Quynh Le, Dingbang Ma, Stanley Perlman, Michael Rosbash

## Abstract

Conventional reverse transcription quantitative polymerase chain reaction (RT-qPCR) technology has struggled to fulfill the unprecedented need for diagnostic testing created by the severe acute respiratory syndrome coronavirus 2 (SARS-CoV-2) pandemic. Complexity and cost hinder access to testing, and long turnaround-time decreases its utility. To ameliorate these issues, we focus on saliva and introduce several advances to colorimetric reverse-transcription loop-mediated isothermal amplification (RT-LAMP) technology; RT-LAMP offers a minimal equipment alternative to RT-qPCR. First, we validated the use of the novel dye LAMPShade Violet (LSV), which improves the visual clarity and contrast of the colorimetric readout. Second, we compared different inactivation conditions on infectivity and RNA yield from saliva. Third, we developed a ten-minute RNA purification protocol from saliva. We call this magnetic bead protocol SalivaBeads. Finally, we developed a magnetic stick, StickLAMP, which provides reliable bead-based RNA purification as well as simple and low-cost access to scalable testing from saliva.

## Introduction

The severe acute respiratory syndrome coronavirus 2 (SARS-CoV-2) pandemic has highlighted many shortcomings in our national response and has driven an enormous increase in our need for in vitro diagnostics. A diagnostic test for SARS-CoV-2 using quantitative real-time polymerase chain reaction (qPCR) technology and nasopharyngeal swabs was developed within weeks of identifying the virus but is suboptimal for serving all diagnostic needs of this global pandemic. This technology has struggled with turnaround time, supply chain shortages, and cost in the effort to increase the amount of testing worldwide^1^. Many entities, commercial and academic alike, have risen to the challenge of adapting and developing molecular technologies to address these shortcomings. Among the myriad efforts that subsequently emerged, reverse transcription loop-mediated isothermal amplification (RT-LAMP) technology may prove to be a viable if not preferable alternative to qPCR technology in addressing at least some of the diagnostic needs of a global pandemic.

LAMP is a nucleic acid detection technology that operates like PCR on the principle of nucleic acid amplification.^2^ Unlike PCR however, LAMP is an isothermal technology and therefore obviates the need for thermal cyclers that would otherwise gate diagnostics behind equipment that costs several thousand dollars. Briefly, LAMP requires at least four (and up to six) different primers: two that contain a self-complementary region that generates a perpetually single-stranded loop structure as well as two that target the region within the single-stranded loop structure. A strand invasion event with the two self-complementing primers initiates production of the initial product containing open loops at either end; exponential amplification can then occur using the loop-targeting primers and a polymerase with strand-displacement ability. In short, LAMP can generate a detectable amount of DNA in a comparable amount of time to PCR but at a single reaction temperature. The DNA can be detected through several different means, including turbidity or through fluorescence with the inclusion of a fluorescent dye. However, a result interpretable to the naked eye would be optimal for a test to enjoy widespread use^3,4^.

Earlier efforts have explored the use of colorimetric detection of amplification products using a pH dye, phenol red, or a magnesium indicator, hydroxynaphthol blue (HNB)^5–9^. With phenol red, the color will change from red to yellow as more DNA acidifies the reaction. With HNB, the color will change from light blue to dark blue in the presence of magnesium which is a byproduct of amplification. These color changes are adequate but suffer from limited visible contrast and can often exhibit ambiguous color changes. Our work here uses the novel pH dye LAMPShade Violet (LSV) as an alternative to phenol red in RT-LAMP^10^. LSV has greater visual contrast between high and low pH and a stronger inflection point, both easing interpretation and reducing the number of ambiguous events.

Saliva is growing increasingly popular as an alternative respiratory specimen to nasopharyngeal swabs for the detection of SARS-CoV-2. Saliva is much easier to collect, and some report that it is a comparable, even superior specimen for SARS-CoV-2 diagnostics^11^. Previous efforts have identified saliva as compatible with RT-LAMP even in the absence of RNA purification. It was replaced by an inactivation step that normalizes the pH, releases RNA, and inactivates RNases^5^. However, saliva is very heterogenous between individuals, which confounds a one-size-fits-all inactivation strategy. We therefore optimized an inactivation protocol with improved success across heterogenous saliva samples when combined with LAMPShade Violet. We suggest that this direct method is suitable for small-scale testing environments where it is easy to resample.

Despite this improvement, many saliva samples were still incompatible with direct input into a colorimetric RT-LAMP reaction. This is prohibitive at scale, where samples cannot be individually resampled or modified for compatibility. We therefore developed and introduced a magnetic bead-based rapid RNA-purification procedure for saliva, which we term “SalivaBeads.” It features a saliva-optimized bead binding solution and uses a magnetic stick (MS) to isolate RNA-bound magnetic beads, which not only improves processing time and scalability but also resolves issues surrounding saliva compatibility with colorimetric RT-LAMP. The SalivaBeads procedure also takes less than ten minutes and substantially improves sensitivity over direct input.

This paper therefore describes our efforts to develop and optimize this low-cost, scalable, and sensitive saliva RT-LAMP protocol, which also minimizes reliance on specialized equipment. The MS is an inexpensive and dramatically more scalable alternative for RNA purification from saliva compared to magnetic racks and multichannel pipettes. We also improved several previous efforts with enhanced visual fidelity, sample compatibility, and sensitivity at minimal additional cost. Altogether, our protocol exhibits a limit of detection of 3.7 copies/µl in a 200µl saliva sample, costs less than $5 per sample without pooling or accounting for labor, and takes approximately 1 hour to conduct from beginning to end.

## Results

We present two different SARS-CoV-2 protocols. The first, named “the direct assay” is conducted on inactivated, unpurified saliva; it is suitable for low throughput testing in low resource environments. However, high frequency or large population testing is likely to prove problematic. This is in part due to the pH variation exhibited by different sources of saliva. In order to address this issue, we developed a second version, “the purified assay”, which adds a novel and rapid purification step. It normalizes saliva pH from different sources while improving sensitivity.

Both protocols begin identically: the crude saliva samples are inactivated by addition of a TCEP, EDTA, and NaOH solution. The samples are then heated to 95°C for 5 minutes and allowed to cool at room temperature for at least 3 minutes. In the direct assay, 5µl of inactivated saliva is then added to two RT-LAMP reactions, one targeting SARS-CoV-2 and the other actin. In the purified assay, 2 volumes of SalivaBeads – described below - are added for 5 minutes before being removed by a magnetic stick. The stick-bound beads are washed in water for 30 seconds and then eluted twice sequentially, first directly into a SARS-CoV-2 RT-LAMP reaction and then into an actin RT-LAMP reaction. In both assays, RT-LAMP reactions are incubated at 65°C for 45 minutes.

Both assays rely on the color difference caused by successful DNA amplification. A positive test is indicated by both reactions turning clear, a negative test is indicated by the actin reaction turning clear and the SARS-CoV-2 reaction remaining purple, and an inconclusive/unsuccessful test is indicated by the Actin reaction remaining purple. Comparing the color changes in the two reactions is critical in the direct assay, where baseline color may be affected by the pH of the input saliva sample. However, a careful comparison is less important in the purified assay, where all samples exhibit comparable baseline pH values following purification.

What follows describes our efforts in developing and optimizing the parameters of our protocol.

### Lamp Shade Violet (LSV) and modifications to avoid heterogeneity to avoid saliva pH variation

In our initial tests, we used a previously described 100x inactivation reagent consisting of 2.5M TCEP, 100mM EDTA, and 1.2M NaOH^5^. The NaOH concentration is critical, as the colorimetric readout varies with pH. As in this publication^5^, we initially used the colorimetric RT-LAMP reagent provided by NEB, which use Phenol Red as a pH sensor. Phenol Red changes from red to yellow upon acidification by successful amplification.

Although 1.2M NaOH was sufficient in most cases, many samples were still too acidic and caused the reaction to turn prematurely positive, meaning even prior to incubation. To address this problem, we took two approaches which were evaluated on four different samples from four individuals (Figure 1A). First, we increased the NaOH concentration to 1.4M and 1.6M and observed the color changes pre- and post-incubation. Second, we used a different pH-sensitive dye – LAMPShade Violet (LSV)^10^. It changes from purple to clear upon successful amplification. Because LSV has sharper contrast and fewer intermediate color changes than Phenol Red, we hypothesized that LSV may help with the interpretation of samples from saliva with outlier pH values.

**Figure 1.**
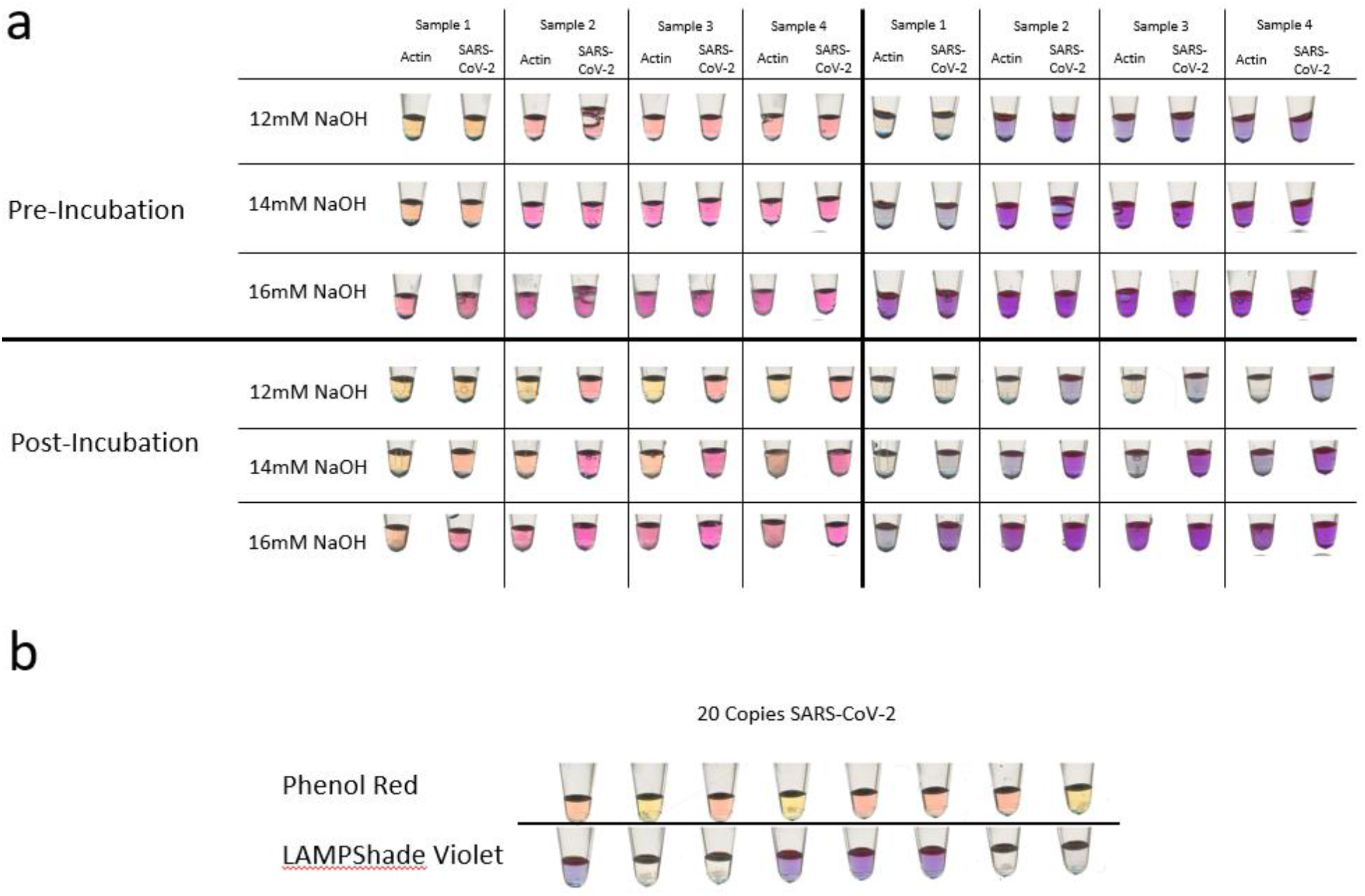
Two comparisons of Phenol Red against LAMPShade Violet for detection of amplification products in RT-LAMP. **(a)** A comparison of color change fidelity and consistency before and after incubation in four varied saliva samples as a function of NaOH. **(b)** A comparison of the sensitivity of the commercial Phenol Red RT-LAMP mix and our in-house reaction mix with LAMPShade Violet judged by their ability to detect 20 total copies of SARS-CoV-2 RNA.

1.4M NaOH was the best concentration to accommodate samples across different pH values. Although a color difference was observed between positive and negative samples, more alkaline samples were still ambiguous with Phenol Red. LAMPShade Violet in contrast was superior in distinguishing positive and negative samples.

To compare our in-house RT-LAMP reaction to NEB-supplied reagents, we used them both to detect 20 copies of SARS-CoV-2 RNA, which is near the threshold for success (Figure 1B). They were qualitatively similar: our in-house version was positive 5 out of 8 times, whereas the commercial was positive 4 out of 8 times. Yet we prefer LSV because of its superior contrast and performance with samples of heterogenous pH values.

### Viral Inactivation at 65°C is effective

The current protocol involves virus inactivation at 95°C followed by a 65°C RT-LAMP incubation. To simplify the protocol and further reduce equipment demands, we assayed virus inactivation at 65°C or even at room temperature (RT) with or without inactivation reagent.

Whereas 5 min at RT was insufficient to eliminate biological activity even with the addition of inactivation reagent, 5 min at 65°C with or without reagent addition reduced activity to undetectable, i.e., by at least 5-6 log units (Figure 2A).

**Figure 2.**
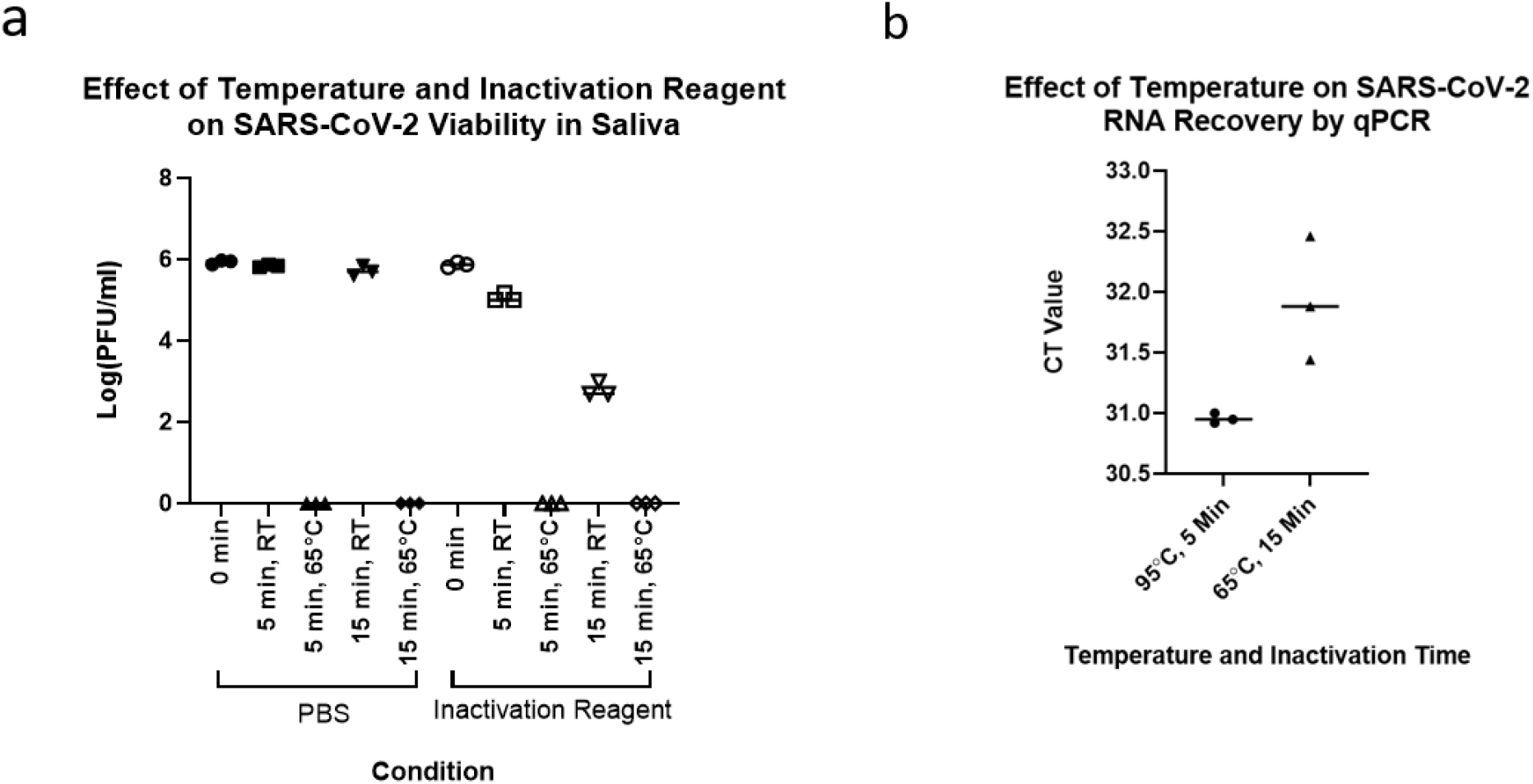
Evaluation of inactivation of saliva samples at 65C. **(a)** A comparison of room temperature and 65C incubations on the viability of SARS-CoV-2 infected Vero E-6 cells with or without inactivation reagent. **(b)** A comparison of RNA recovery from encapsulated inactivated SARS-CoV-2 virions in saliva at 95C for 5 minutes and 65C for 10 minutes.

We also compared SARS-CoV-2 RNA yield from a 65°C vs a 95°C incubation. To create specimens that most closely resembled clinical samples, we contrived saliva samples using inactivated but intact SARS-CoV-2 virions from Zeptometrix (Figure 2B). A 65°C degree inactivation for 15 minutes led to a significant 1-cycle, approximately 2-fold loss in RNA yield compared to a 95°C inactivation for 5 min (t(4)=3.274, p=0.0307). This modest decrease suggests that 65°C inactivation is an acceptable alternative in environments that can only afford minimal equipment or are otherwise averse to near-boiling temperatures. However, the improvement in yield at 95°C suggests that it is preferred in environments that can accommodate this temperature,. Further efforts described below use this inactivation temperature.

### Testing means of purifying RNA from saliva

To further improve sensitivity and sample compatibility, we sought to develop a rapid RNA purification protocol optimized for saliva. We first measured RNA recovery using fluorimetry with a Qubit device and compared various bead-based methods with Trizol purification (Figure 3A). They included: 2 volumes of Ampure Beads XP, 2 volumes of magnetic silica beads in an NaCl/PEG-8000 solution, 2 volumes of magnetic carboxylated beads in an NaCl/PEG-8000 solution, glass milk in a NaCl/PEG-8000 solution as well as glass milk in a Sodium Iodide (NaI) solution as previously described^5,12^. Ampure XP beads as well as the NaCl/PEG-8000 based bead mixes were comparable to Trizol (Figure 3A). Due to lower cost and the advantage of being suitable for scale and automation, we decided to use the NaCl/PEG-8000 based bead mix with carboxylated beads for further optimization of RNA purification from saliva.

**Figure 3.**
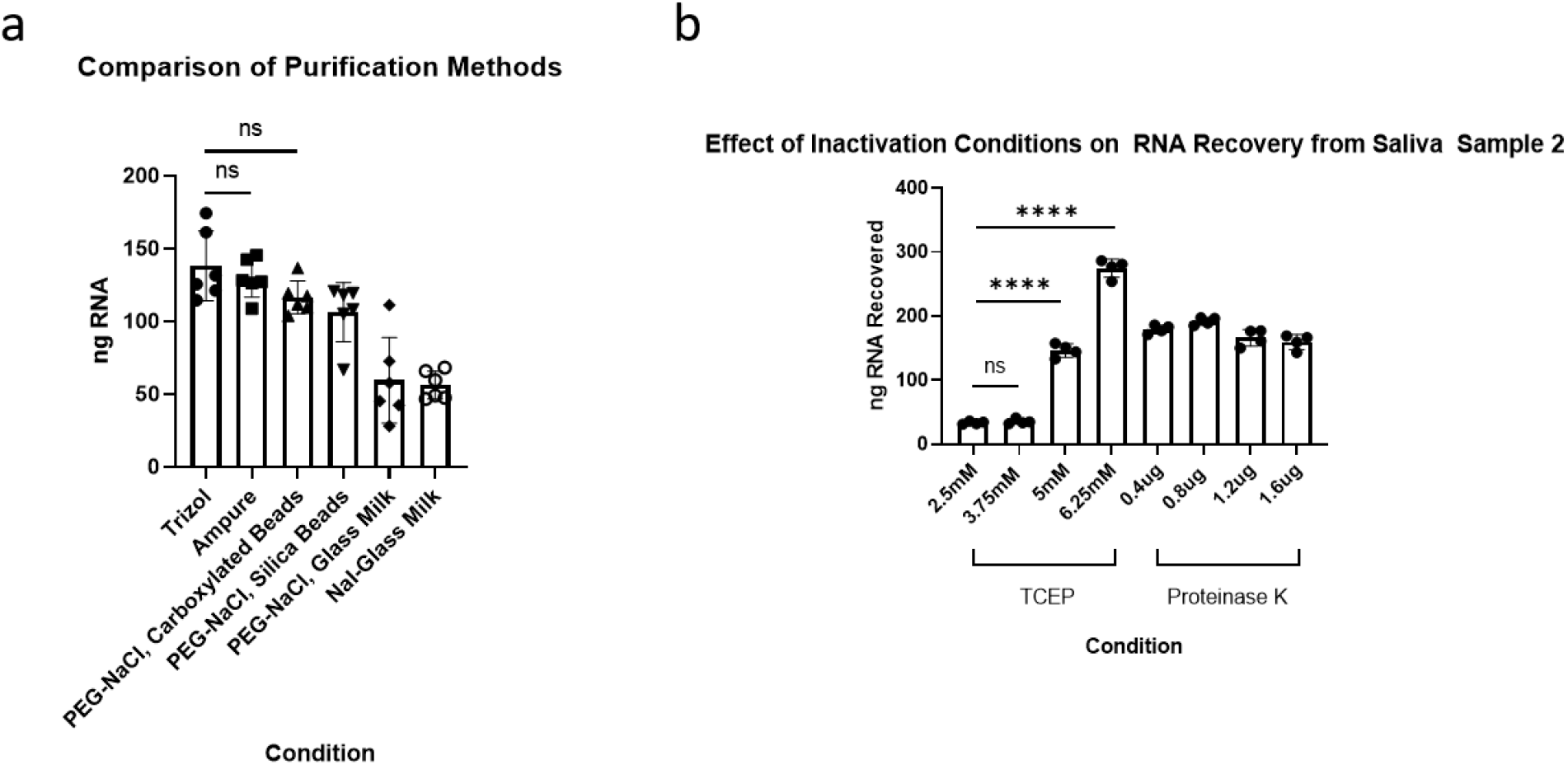
Investigating and optimizing saliva for purification. **(a)** A comparison of select purification methods on total RNA recovered from saliva measured by Qubit fluorimetry. **(b)** A comparison of RNA recovered from PEG-NaCl, Carboxylated beads following inactivation methods using different concentrations of TCEP or Proteinase K.

Because current inactivation procedures were developed for direct input of saliva into an RT-LAMP reaction, we further optimized inactivation for input into bead-based RNA purification. Different concentrations of TCEP as well as Proteinase K were assayed, the later with a 5 min 65°C incubation followed by a 5 min 95°C incubation (Figure 3C).

Although Proteinase K was effective in increasing RNA yield compared to the initial condition of 2.5mM TCEP, 6.25mM TCEP was even better and used below.

### Optimizing salt and salt concentration

To optimize the NaCl/Peg-8000 bead mix, we examined the effects of salt type, salt concentration, and PEG-8000 concentration on RNA yield from saliva. For reference, the original purification recipe calls for 1M NaCl, and 18% PEG-8000^12^. Different concentrations of NaCl and different concentrations of Sodium Acetate (NaOAc) were assayed; the latter is also commonly used in nucleic acid precipitation. Different concentrations of Guanidine Hydrochloride (GuHCl) – another reagent commonly used in nucleic acid purification – were also assessed (Figure 4A). Although all salts at all concentrations tested could purify RNA, NaCl at a reduced concentration of 0.7M produced a significant higher yield compared to the original recipe at 1M (P<1×10^−4^, One-Way ANOVA, Tukey’s HSD) and compared to the other salts.

**Figure 4.**
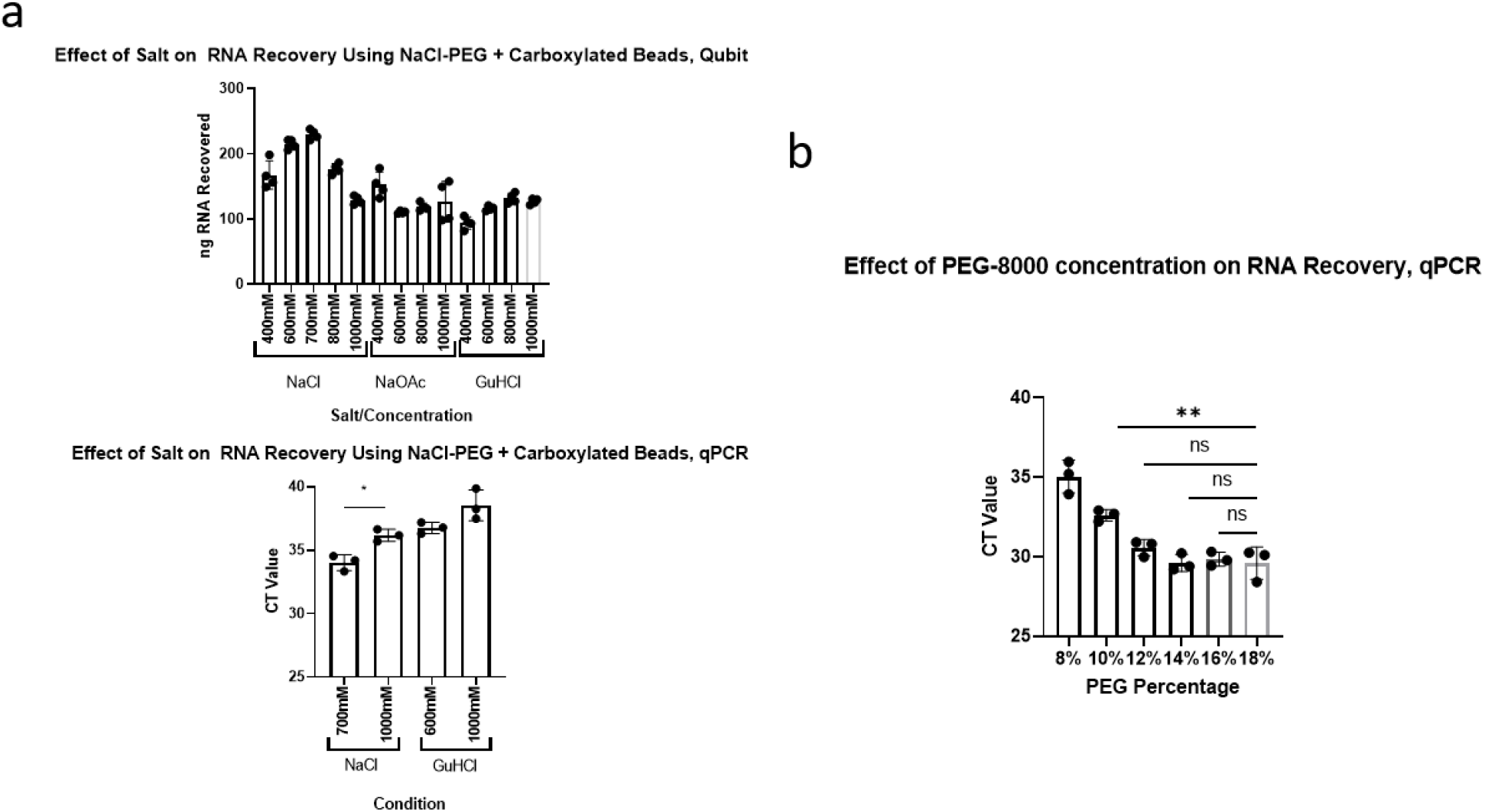
Optimizing the concentrations of salt and PEG for RNA purification from saliva. **(a)** A Comparison of multiple salts at multiple concentrations on RNA recovery from Saliva with 18% PEG. **Top:** Measurement of recovery by Qubit fluorimetry. **Bottom:** Measurement of SARS-CoV-2 RNA recovery by N1 qCPR from contrived saliva samples. **(b)** A comparison of the effect of multiple PEG concentrations with 700mM NaCl on RNA recovery, measured by SARS-CoV-2 RNA recovery by N1 qPCR from contrived saliva samples.

In other tests, the addition of PEG-8000 to RT-LAMP increased the false positive rate (data not shown). Although we did not see a similar change with the purified assay, it is possible that a modestly increased rate becomes relevant with large scale testing. We therefore decided to reduce the concentration of PEG-8000 until it affected yield. We found no significant difference between 18% PEG-8000 and 12% PEG-8000 (Figure 4B). Because visual examination of the data suggested a slight loss of sensitivity between 14% and 12% (data not shown), we adjusted the buffer to 14% PEG-8000. Our final adjusted recipe therefore uses 0.7M NaCl and 14% PEG-8000.

### Optimizing elution and binding times

In our purification protocol, the bead mix is left to bind RNA for a certain amount of time before being removed by a magnet, henceforth referred to as binding time. At the end of the protocol, the beads are left to release RNA in the reaction mix for a certain amount of time, henceforth referred to as elution time. Our earlier development procedures used a 5 minute binding time and a 1 minute elution time. To minimize the test processing time, we determined the minimal binding and elution times without sacrificing sensitivity.

We first tested a 1 minute, 5 minute, and 15 minute binding time with a 1 minute elution. We observed a significant difference between 1 minute and a 5 minute binding time (p < 0.01, One-Way ANOVA, Tukey’s HSD) but no significant difference between 5 minutes and 15 minutes (Figure 5A). We therefore chose a 5 minute binding time. We then compared 10 second, 1 minute, and 5 minute elution time (Figure 5B). We found no significant differences and so chose an elution time of 30 seconds for improved operational consistency.

**Figure 5.**
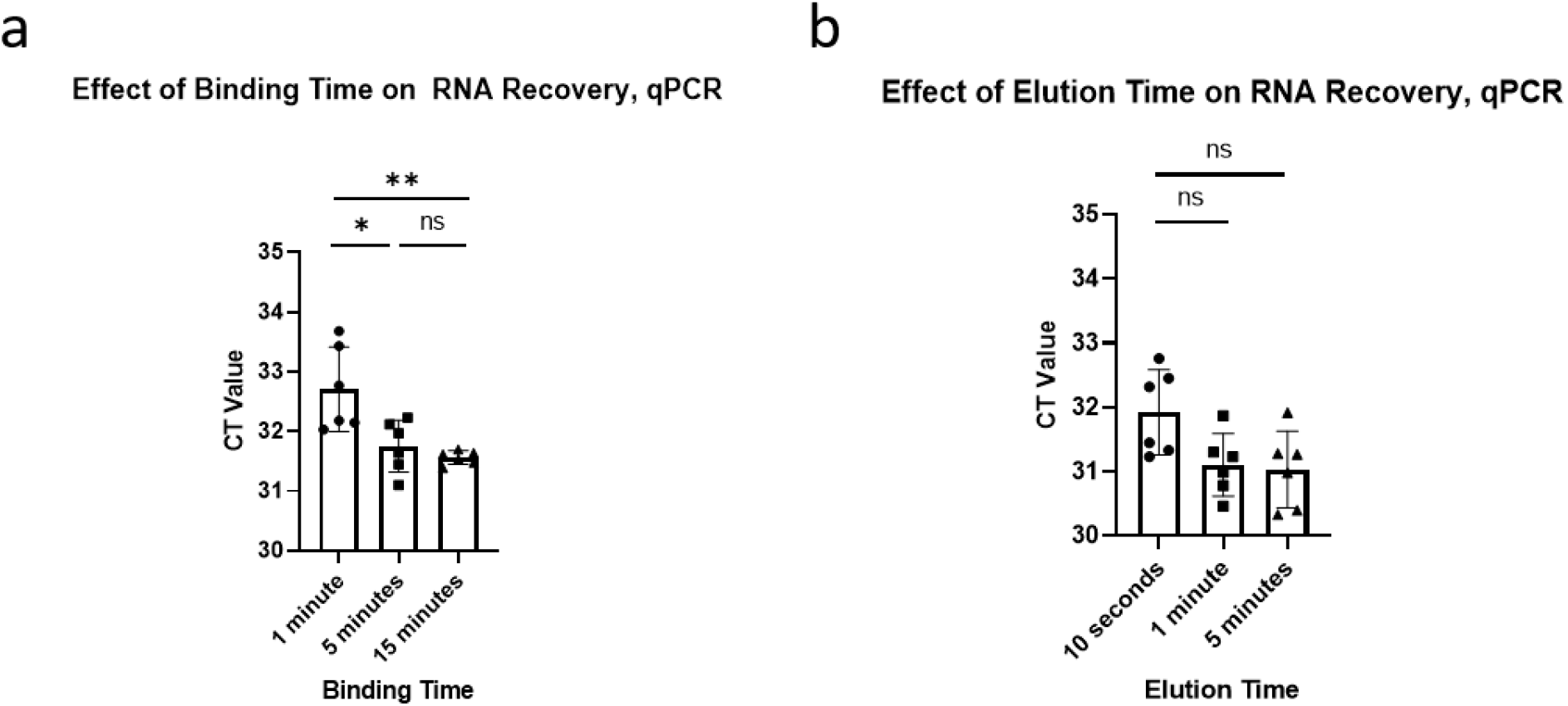
Optimizing the binding and elution times for magnetic bead-based purification using carboxylated beads with 700mM NaCl and 14% PEG, as measured by SARS-CoV-2 RNA recovery through N1 qPCR from contrived saliva samples. **(a)** The effect of binding time on RNA recovery, with a 1-minute elution. **(b)** The effect of elution time on RNA recovery, with a 5-minute binding time).

**Figure 6.**
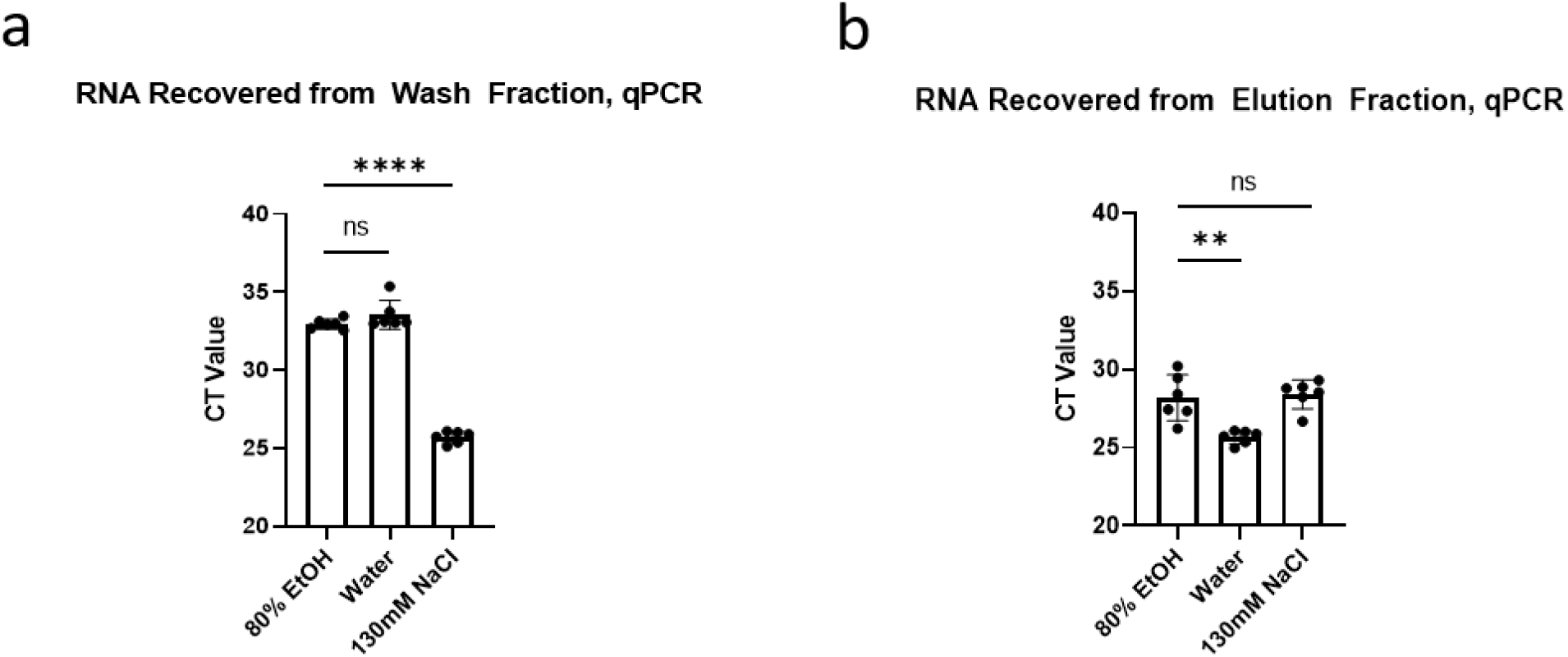
A comparison of three washing conditions for RNA purification from saliva using carboxylated beads with 700mM NaCl and 14% PEG, as measured by SARS-CoV-2 RNA recovery by N1 qPCR from contrived saliva samples. **(a)** A measurement of RNA recovered from a 30 second wash in the indicated wash buffer. **(b)** A measurement of RNA recovered from a 30 second elution in water from samples washed using the indicated wash buffer.

### Optimizing wash buffers

The majority of bead-based RNA purification protocols feature a 70-80% ethanol wash to remove non-specifically bound contaminants without removing nucleic acids. Because ethanol is incompatible with many downstream enzymatic reactions, protocols involving an ethanol wash typically require a long drying step to remove all traces of ethanol. To eliminate this drying step, we compared this standard 80% ethanol wash to Water and to 130mM NaCl, which was used in a cellulose dipstick-based purification assay.^13^ We therefore washed the beads for 30 seconds in 200µl of the indicated wash buffers and then eluted in 50µl of Deionized H2O for 30 seconds. We then ethanol precipitated the RNA under standard conditions (see methods) and assayed recovery by qPCR.

There was no significant difference between RNA levels in the 80% EtOH and in the Water wash fractions (Figure 7A). However, 130mM NaCl washed off significantly more RNA (p<1×10^−4^, One-Way ANOVA, Tukey’s HSD). In the eluate, there was no significant difference between RNA recovered from the 130mM NaCl-washed beads and the 80% EtoH-washed beads, but the H2O-washed beads exhibited a significant 1-cycle improvement (p<1×10^−3^, One-way ANOVA, Tukey’s HSD) (Figure 7C). We therefore decided that water was the most suitable wash reagent for quick purification, also because water is compatible with the RT-LAMP reaction and requires no drying step following washing.

**Figure 7.**
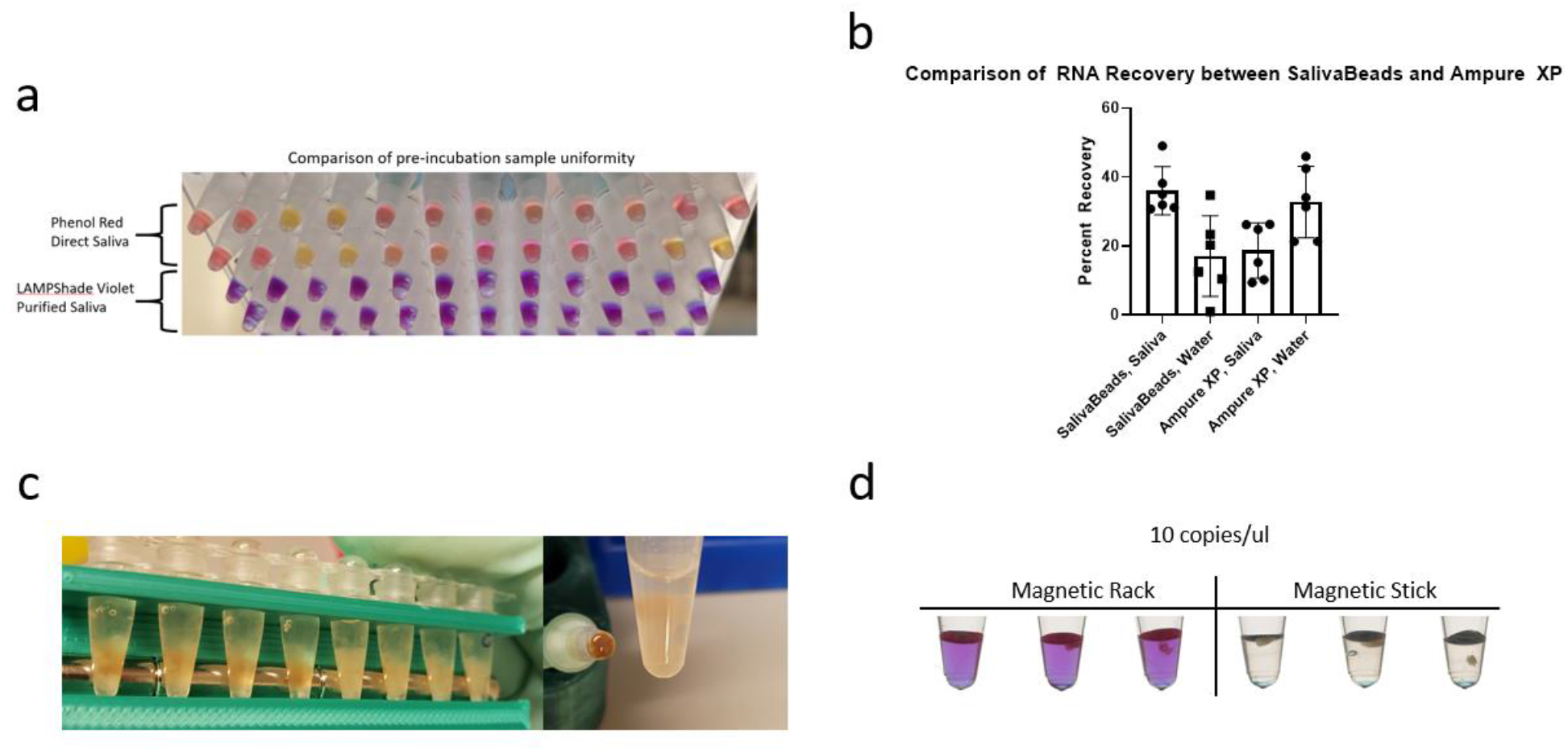
Several features of SalivaBeads purification. **(a)** A comparison of pre-incubation sample color uniformity in 12 samples, with SARS-CoV-2 and Actin reactions side by side. Direct input of inactivated saliva in a commercial Phenol Red-based reaction is compared to a SalivaBeads-purified RNA input into our LAMPShade Violet-based reaction. **(b)** A comparison of SARS-CoV-2 RNA recovered from contrived saliva samples by N1 qPCR, measured by number of copies recovered by qPCR divided by number of copies contrived into saliva. **(c)** A visual comparison of debris bound to magnetic-beads when isolating beads by magnetic rack (left) or a magnetic stick (right). **(d)** A comparison of sensitivity between SalivaBeads purified via magnetic rack (left) and magnetic stick (right), by RT-LAMP with LAMPShade Violet.

These changes and optimizations and our bead mix are henceforth referred to as “SalivaBeads”.

### StickLAMP

Most nucleic acid isolation procedures that use magnetic beads also employ an elution step, which releases the purified RNA into an intermediate buffer such as water or Tris prior to addition to a reaction. We sought to simplify this strategy by eluting RNA from SalivaBeads directly into the RT-LAMP mix. However, the sensitivity was poor compared to the direct assay. Both assays were able to routinely detect 100 copies/µl of SARS-CoV-2 RNA, but only the direct assay was able to detect 25 copies/µl. (Figure 7A).

To address whether this was due to a poor RNA yield from SalivaBeads, we compared this yield to that from Ampure XP beads. This was done from saliva as well as from purified RNA in water (Figure 7B). SalivaBeads had superior yield to Ampure XP beads from saliva but inferior from water. Moreover, the SalivaBeads yield from saliva was comparable to the Ampure XP yield from water. This indicates that the reduced sensitivity is not due to poor yield but rather from some sort of contaminant carryover or other incompatibility.

Indeed, we noticed that considerable debris clung to the SalivaBeads that persisted through the wash step and contaminated the RT-LAMP mix (Figure 7C, left). We therefore developed a magnetic stick that would remove the beads prior to downstream processing (Figure 7D). Importantly, the stick appeared to be selective for removing the beads without most of the saliva debris (Figure 7C, right). Consistent with this observation, magnetic stick purification could faithfully detect 10 copies/µl, whereas magnetic rack purification could not (Figure 7E).

### Performance analysis of StickLAMP

To evaluate the performance of StickLAMP purification, we determined the copy number at which 95% of reactions score positive, henceforth referred to as the limit of detection (LOD). Our LOD was at least 3.7 copies per microliter, i.e., 19 of 20 contrived samples with 3.7 copies of SARS-CoV-2/µl in 200ul saliva scored positive (Figure 8A).

**Figure 8.**
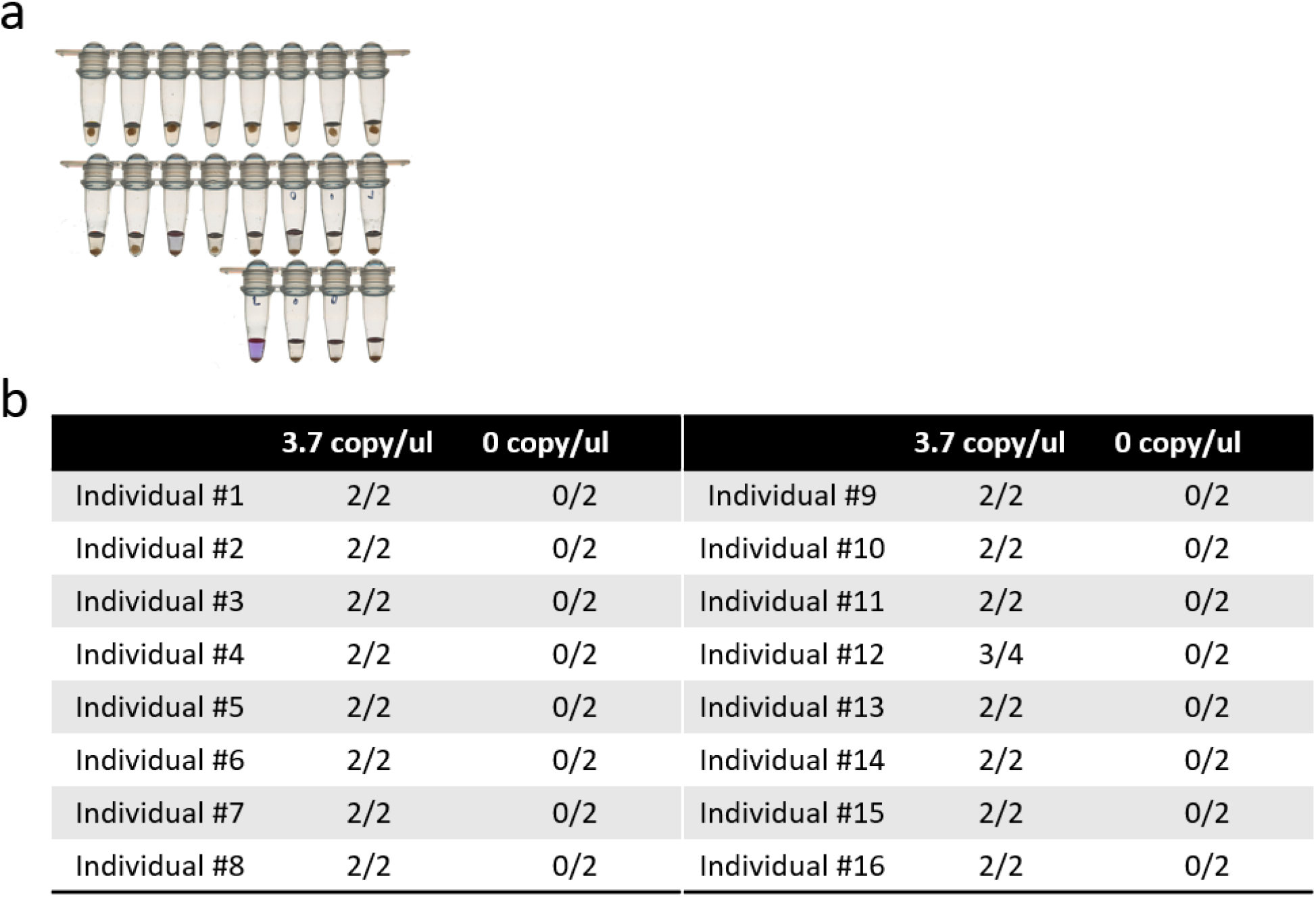
Performance evaluation of SalivaBeads and StickLAMP. **(a)** Limit of detection experiment: Ability of StickLAMP to detect 3.7 copies/µl of SARS-CoV-2 RNA from 200µl of contrived saliva in 20 replicates. **(b)** Testing the ability of StickLAMP to detect 3.7 copies/µl in 200µl saliva from 16 different samples.

To ensure that this LOD is similar across a wide variety of saliva types, we collected saliva from 16 different individuals and created two contrived samples per individual with 3.7 copies/µl of SARS-CoV-2 (Figure 8B). For 15/16 individuals, both samples were positive. For the 16^th^ individual, only one sample was positive, but both samples were positive upon retesting. The 3.7 copy/µl LOD is therefore robust across different individuals.

## Discussion

We present two protocols for the detection of SARS-CoV-2 RNA from saliva, which feature several innovations. First, LAMPShade Violet (LSV) is an attractive alternative to Phenol Red for the colorimetric detection of SARS-CoV-2 RNA in saliva; LSV improves visual fidelity without sacrificing sensitivity. Second, 65° is completely successful at viral inactivation in saliva. Compared to the more standard 95°C, 65°C enhances safety, reduces testing time and equipment demands with only somewhat reduced detection sensitivity. Third, we present a rapid purification protocol. It adds less than ten minutes to the processing time, costs less than 20 cents per sample, and is optimized for saliva. The purification improves sensitivity over tenfold and normalizes sample pH for downstream colorimetric detection.

Purification uses a magnetic stick (MS)as an integral tool for rapid nucleic acid purification. Saliva contains substantial and variable levels of debris. It sticks to nucleic acid binding media like beads and inhibits the LAMP colorimetric assay. One previous effort also focused on this assay circumvented the debris issue with centrifugation^5^. However, this solution is undesirable for scaled testing as centrifuges are difficult to introduce into an automated workflow. They are also expensive and are contrary to the goal of minimizing equipment requirements. The MS selectively binds magnetic beads over saliva debris and therefore substitutes for centrifugation. MS also has several advantages over the other traditional method of bead separation, magnetic rack purification.

First, MS has superior scaling potential. Multichannel pipettes are typically used in magnetic rack purification to improve throughput and sample processing time in both manual and automated workflows. Importantly however, the number of channels is constrained by mechanical considerations in multichannel pipettes, whereas the MS simplicity enables the number of simultaneous “channels” to scale well beyond traditional limitations. Indeed, we developed 4-channel, 8-channel, and a 24-channel versions for the purpose of large-scale multiplex purification reactions, as well as a custom rack designed for a 24-channel MS to elute directly into a 96 or 384-well plate (Figure 9B and 9C).

**Figure 9.**
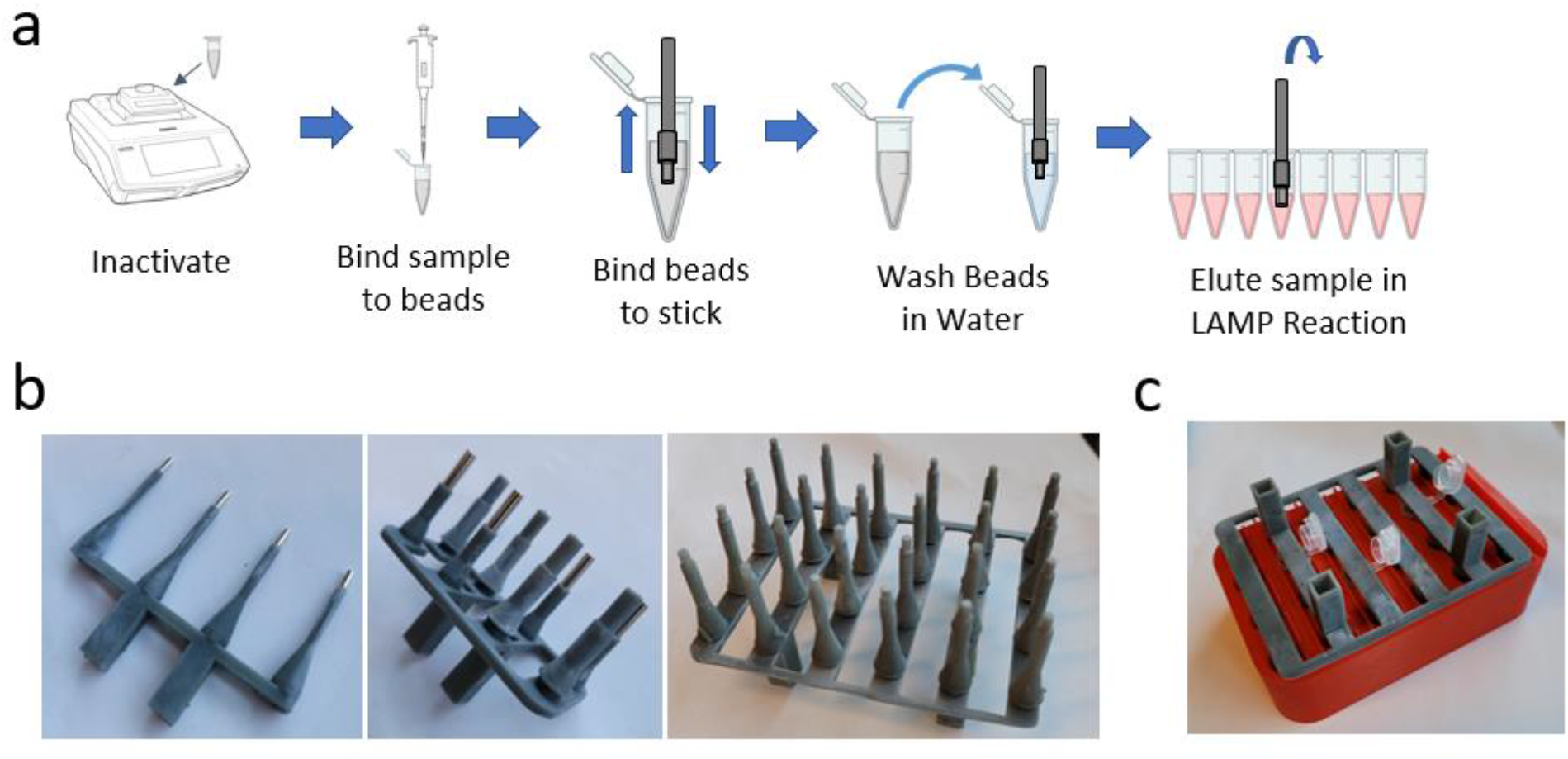
An overview of StickLAMP and further considerations. **(a)** A visual overview of the StickLAMP protocol. **(b)** A 4-channel magnetic stick (Left), 8-channel (Middle), and 24-channel (Right). **(c)** A demonstration of a 24-channel magnetic stick in 3D-printed hardware designed to hold 24 1.5ml Eppendorf tubes.

Second, this superior scaling potential also reduces the additional processing time required by more samples. The addition of every 8 samples increases the required processing time with an 8-channel multipipette, but a 24-channel MS increases processing time only beyond 24 samples. The processing time is further reduced because 2-3 pipetting steps are replaced with a single dipping step.

Third, the cost of a MS is dramatically less than a multichannel pipette. A 24-channel magnetic stick costs less than $5.00 to produce, and each tip is less than $0.05, about $6.00 in total. An 8-channel multipipette in contrast costs well over a thousand dollars.

Our purification optimization results also suggest several unexplored directions for future saliva-based nucleic acid diagnostics. The optimal NaCl concentrations were well below the theoretical limit required for purifying nucleic acids, suggesting that uncharacterized minerals or salts present in saliva are aiding nucleic acid binding to carboxylated beads^14^. In addition, all saliva samples benefitted from additional TCEP, greater than previously published concentrations^5^. However, the degree to which they benefit and even the amount of RNA purified from different saliva samples varied. This suggests potential heterogeneity of RNase levels and/or RNA content between samples, which future saliva-based diagnostic test efforts should consider to make further improvements. We are nonetheless confident in our reported LOD, given its consistency across the wide variety of tested samples.

In summary, these new SARS-CoV-2 detection protocols are inexpensive, less than $5 per test without considering labor and sample pooling, and offer improved scalability over existing tests without sacrificing sensitivity. They are especially suited for workplaces or schools with modest numbers of employees and students, from single digits to the low thousands. The minimal equipment requirements and low cost also make them well-suited for low resource environments, which still might be able to mount a medium complexity CLIA lab. We note in this context that low-resource and underserved environments have been disproportionately vulnerable to the spread of SARS-CoV-2^15^.

## Data Availability

All data is reported in the manuscript. Raw data is available upon request.

## Acknowledgements

This work was supported by the Howard Hughes Medical Institute. We express our thanks to the gLAMP consortium for their willingness to create an open dialogue and engaging in helpful discussions and ideas in an exceptionally collaborative space. We would like to thank Brian Rabe and Connie Cepko of Harvard Medical School for sharing the reagents and ideas that initiated this project. We also express enormous gratitude to Tim Brown, Luke Lavis, and Ronald D. Vale for sharing LAMPShade Violet with us, as well helping us through constructive dialogue and general camaraderie. We would like to thank Kate Abruzzi for reading, editing, and generally helpful discussions. Finally, we would like to thank the Brandeis facilities staff for helping facilitate this endeavor.

## Methods

### Oligonucleotides

#### RT-LAMP Reactions

Phenol Red experiments were conducted using WarmStart Colorimetric LAMP 2x Master Mix (NEB, M1800L) in 25µl reactions with 5µl of inactivated saliva. Lampshade violet experiments were conducted using a buffer consisting of 5mM Tris ph8.5, 8mM MgSO4, 30mM KCl, 0.1% Tween-20, 10mM dNTPs, 12.5mM KOH, 10mM LampShade Violet^10^(Luke Lavis, Janelia Research Labs), 0.5µl WS RTx (NEB M0380L), WS BST 2.0 (NEB M0538L) in 25µl reactions with 5µ of inactivated saliva. For magnetic stick experiments, SARS-CoV-2 reactions were conducted in 25µl reactions, while Actin reactions were conducted in 20µl. All reactions were heated to 65°C for 45 minutes and cooled to at least room temperature prior to examination.

Reactions were imaged on an Epson V850 Scanner.

#### Primers

**Table 1.**
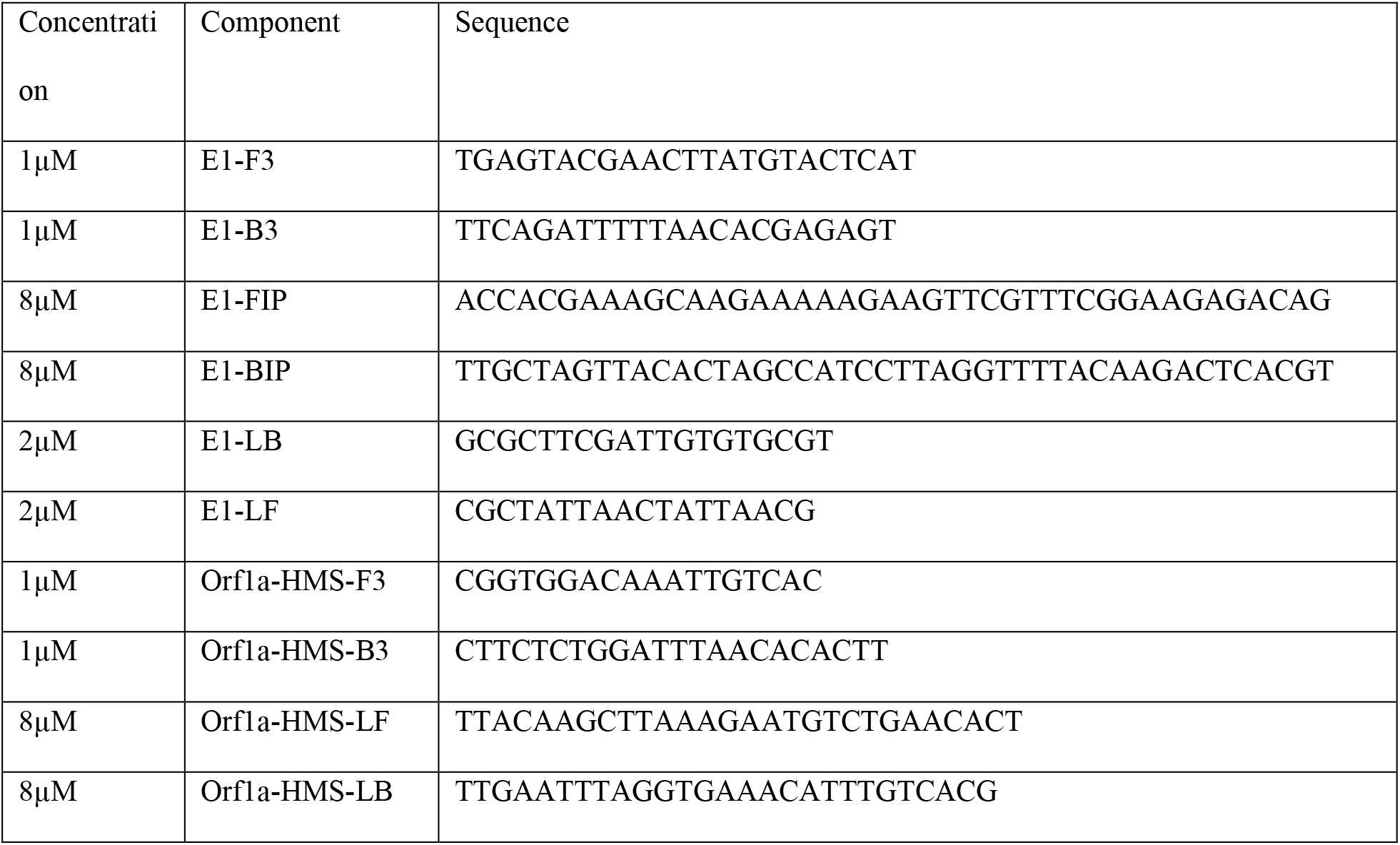
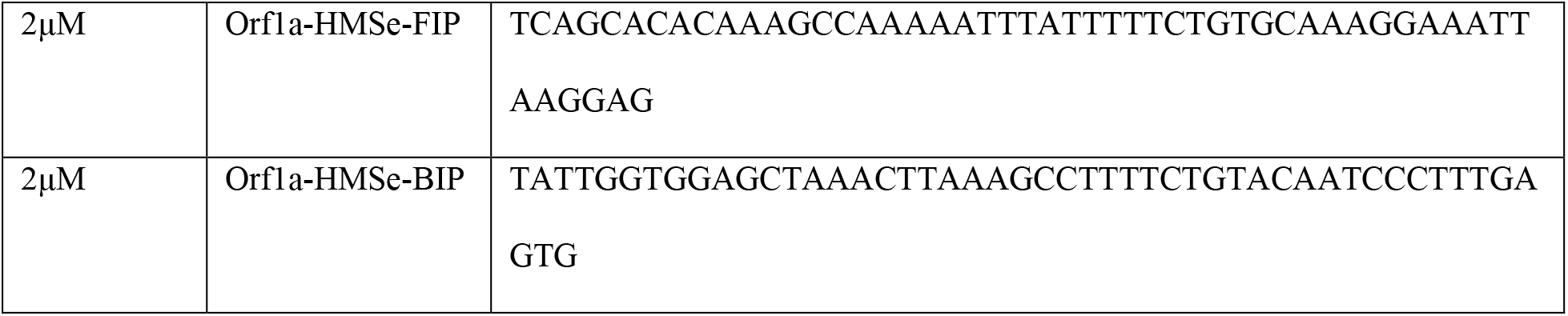
describes primer sequences and concentrations used to produce a 5x primer solution. Primer sequences are derived from previous work^4,5^.

#### Control RNA

Contrived samples were created using either heat-inactivated SARS-CoV-2 RNA from BEI (NR-52286), or inactivated SARS-CoV-2 viral particles from Zeptometrix (CAT# NATSARS(COV2)-ST). In order to optimize and evaluate magnetic-bead purification procedures, RNA from BEI was diluted in 1ng/µl Drosophila RNA to 1000 copies/µl and spiked in to inactivated saliva at the stated concentration. In order to evaluate the full-process performance of the protocol, limit of detection experiments and experiments intended to evaluate RNA release from viral particles were conducted using Zeptometrix SARS-CoV-2 particles added into raw saliva at the stated concentrations. In the proceeding methods, the control used will be specified.

#### Saliva inactivation

10x inactivation was prepared with 62.5mM TCEP(Goldbio TCEP1), 10mM EDTA (ThermoFisher Scientific 10977015), and 130mM NaOH. 10x inactivation reagent was added to a final concentration of 1x to raw saliva and inverted 10 times, or vortexed for 5 seconds at maximum speed. Saliva samples were then heated to 95°C for 5 minutes and allowed to cool at room temperature for 3 minutes prior to downstream processing. When testing 65°C inactivation, we inactivated for 15 minutes at 65°C and let samples cool to room temperature for 3 minutes prior to downstream processing.[But what about 65 degree change?]During optimization experiments, TCEP concentrations used are as described in figures and results.During Proteinase K testing, the indicated amount of Proteinase K (NEB P8107S) was added to saliva and heated at 65°C for 5 minutes and inactivated at 95°C for 10 minutes. Virus titers in untreated or treated samples were then determined using Vero E6 cells (grown in 10% FBS-DMEM). For plaque assays, cells were fixed with 10% formaldehyde 3 days after infection and stained with crystal violet.

#### Purification

Commercial Ampure XP beads (Beckman-Coulter Life Sciences A63881) were used according to protocol at 2x volumes and eluted in 30µl Milli-Q H_2_O.

Unmodified homemade magnetic bead buffer was prepared using 100mM NaCl (Sigma-Aldrich S9888), 20% PEG-8000 w/v (Sigma-Aldrich 1546605), 10mM Tris-HCl, pH 8, 1mM EDTA (ThermoFisher Scientific 10977015, and Milli-Q H_2_O to 50ml. 1000µl Sera-mag Speedbeads (FisherScientific 09-981-123) were washed twice in 1ml 10mM Tris ph 8, 1mM EDTA, and added to bead buffer.

The final SalivaBeads recipe uses the same materials as listed above, but with 700mM NaCl, 14% PEG-8000 w/v, 10mM Tris-HCl, pH8, 1mM EDTA, 200µl Sera-mag Speedbeads, and Milli-Q H_2_O to 50ml.

Commercial Ampure XP beads and unmodified homemade magnetic bead buffer was used according to commercial Ampure XP protocols.

For Salivabeads, two volumes of SalivaBeads were added to inactivated saliva samples and inverted 5 times. Bead+saliva mix was incubated at room temperature for 3 minutes. A magnetic stick with tip was added to bead+saliva mix for two minutes, with agitation at one minute and before removing. Magnetic stick with bound beads were dipped in 100µl Milli-Q H_2_O prepared in a PCR strip tube up and down 5 times, then left to incubate for 20-25 seconds. The magnetic stick was then removed from the water and placed in SARS-CoV-2 reaction mix for 60 seconds. After 60 seconds, the magnetic stick was removed from the SARS-CoV-2 reaction mix and placed in Actin reaction mix for 30 seconds, then removed and the tip was discarded.

For purifying RNA from wash steps, ethanol was added to each sample to a final concentration of 75% and a final volume of 200µl. Sodium acetate (Invitrogen AM9740) was added to a final concentration of 0.3M. Samples were left to precipitate overnight at -20°C. The next day, samples were spun down for 30 minutes at 18000 RCF. Samples were washed once in 80% ethanol and eluted into 20µl of Milli-Q H_2_O, 5µl of which was subsequently used in qPCR reactions.

#### qPCR

qPCR was done using Luna Universal Probe One-Step RT-qPCR Kit (NEB E3006L) and the CDC N1 primer (IDT 10006713). Reactions and cycling conditions were prepared according to manufacturer’s protocol.

#### 3D Printing

Magnetic Sticks and Tips were printed using Siraya Tech Blu (Siraya Tech) on an Epax X10 UV LCD 3D Printer with 8.9 inch 4K mono LCD (Epax) using the following settings: 0.05mm Layer Height, 8 Bottom Layers, 3.2s Exposure Time, 12.4s Bottom Exposure Time, 7mm Lift, 35mm/min Lift Speed, 125mm/min Retract Speed. Magnets used were 2.54mm Diameter, 0.600” Length N50 magnets (SuperMagnetMan, Cyl0072-20).

